# The MagMa Study: Quantum Magnetocardiography in Non-Ischemic Cardiomyopathy

**DOI:** 10.1101/2025.05.29.25328567

**Authors:** Phillip Suwalski, Finn Wilke, Ekaterina Latinova, Gabriele Paci, Gamze Satilmis, Karin Klingel, Sebastian Kelle, January Weiner, Dieter Beule, Thomas F. Lüscher, Landmesser, Bettina Heidecker

**Author notes:** **Corresponding author:** Bettina Heidecker, MD, Deutsches Herzzentrum der Charité (DHZC), Campus Benjamin Franklin, Department of Cardiology, Hindenburgdamm 30, 12203 Berlin, DECT.

## Abstract

**Background:** Rapid diagnostic screening is an unmet need in patients with suspected cardiomyopathy. We recently demonstrated that magnetocardiography (MCG) might be a suitable tool to detect cardiomyopathies. We now tested diagnostic accuracy of MCG prospectively.

**Methods:** MCG uses a superconducting quantum interference device (SQUID) to detect the electromagnetic field of the heart. In our recent retrospective study, a T-beg-Tmax (MCG Score) > 0.051 identified patients with non-ischemic cardiomyopathy. We assessed the diagnostic accuracy of MCG compared to advanced imaging and/or endomyocardial biopsy (EMB). Results from EMB took precedence over advanced imaging. We enrolled 110 patients with angina-like symptoms after exclusion of coronary artery disease (Power calculation: Expected 80% power at a 5% significance level) from the emergency department, in-patient wards and outpatient clinics. Exclusion criteria included presence of intracardiac metal devices and treatment with immunosuppressants. We incorporated 220 healthy individuals using 1:2 propensity score matching (age, sex) for a total of 330 participants.

**Results:** MCG detected cardiomyopathy with 94.74% sensitivity (95% CI: 85-99%), 98.54% specificity (95% CI: 96-100%), 93.1% negative predictive value (NPV, 95% CI: 84-98%) and 94.34% positive predictive value (PPV, 95% CI: 84-98%). In 3 patients, CMR failed to detect non-ischemic cardiomyopathy, whereas MCG showed pathological results consistent with results of EMB. Results were highly reproducible (statistical uncertainty: 2.5%, T-beg-Tmax (MCG Score) +/− 0.004).

**Conclusion:** MCG offers a non-invasive, reproducible methodology for rapid and accurate detection of non-ischemic cardiomyopathy. Our data indicate that MCG may be an efficient tool for allocation of advanced imaging to those in greatest need.

**Visual Abstract:** 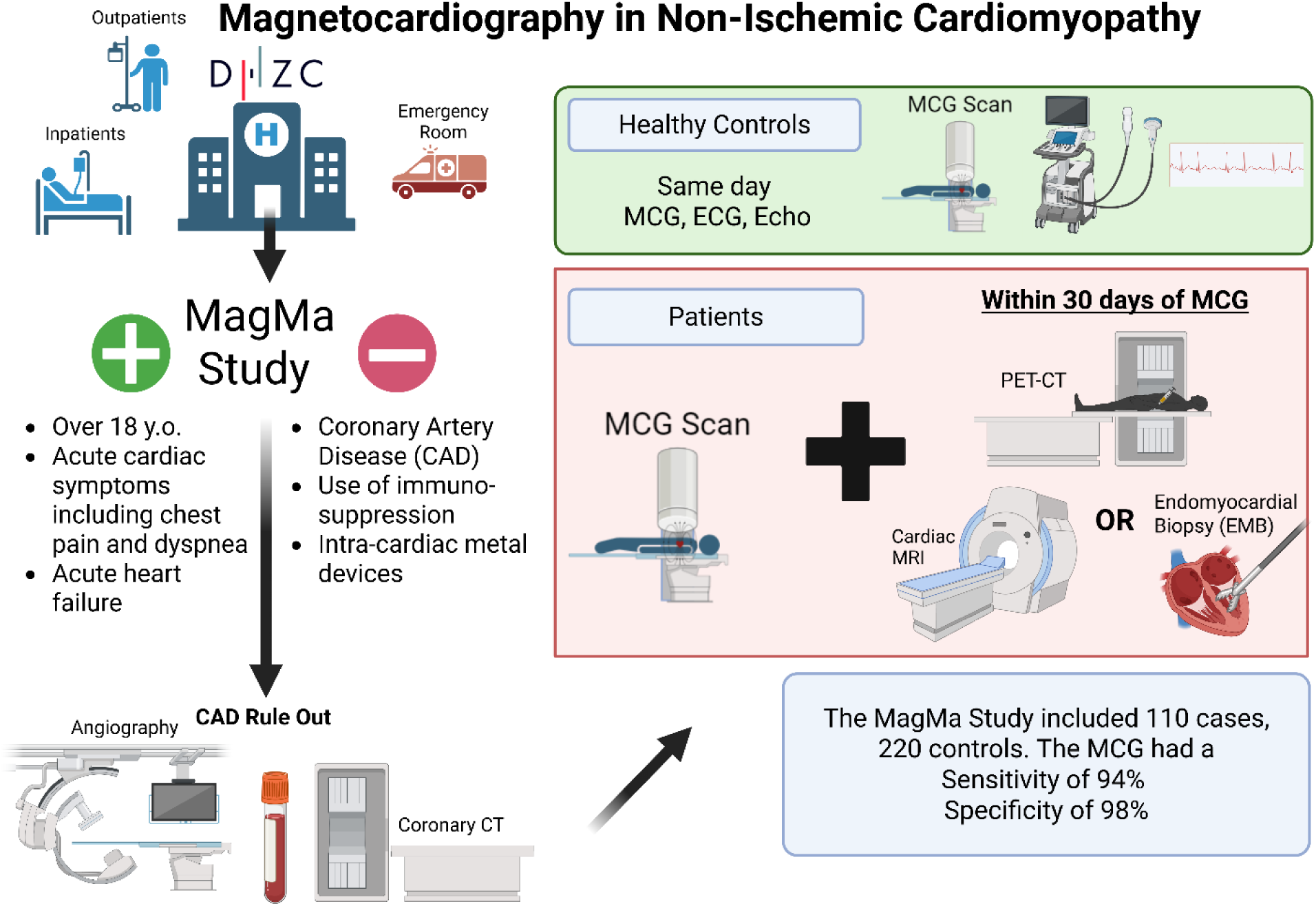

## Introduction

Accurate and rapid screening is an unmet medical need to address the increasing demand for advanced diagnostic workup in patients with suspected cardiomyopathy (1–3). In patients with chest pain or angina-like symptoms in whom coronary artery disease has been ruled out, advanced imaging is recommended by the guidelines American Heart Association (Class 1) (4) and the of the European Society of Cardiology (Class 1) (5, 6) to evaluate for cardiomyopathies. Due to an at times overwhelmed healthcare system, advanced imaging such as cardiac magnetic resonance imaging (CMR) is not always readily available, which can result in underdiagnosis and turn progression of cardiomyopathies to advanced heart failure (7, 8). Advanced heart failure is associated with an increased risk of sudden cardiac death and may require treatment with mechanical circulatory support or even heart transplantation (9). We recently demonstrated in a retrospective study that magnetocardiography (MCG) may be a suitable tool to detect non-ischemic cardiomyopathies in a highly effective manner without any safety concerns (10–12). The feasibility of using MCG for diagnosing non-ischemic cardiomyopathies has since been confirmed by an independent group employing a different device, underscoring both the robustness of the findings and their reproducibility across platforms. (13).

MCG uses quantum technology to detect the electromagnetic field of the heart. The electromagnetic field is created by the activity of ion channels of the myocardium. Similar to the electric field that is measured by an ECG, also an electromagnetic field is created, which can be measured through MCG. Our retrospective study demonstrated that his electromagnetic field changes as cardiomyopathy develops (14, 15). As a non-invasive technique, MCG offers distinct advantages for screening and monitoring of heart disease. First, MCG measurements are acquired without contact in just 60 seconds (15). Second, it is widely applicable to most patients, with the exception of those with metal implants in the chest, which may interfere with magnetic field detection (15). Third, MCG generates an objective score, minimizing interobserver variability and ensuring high reproducibility. Lastly, MCG provides unparalleled precision in detecting even small changes in the magnetic field (10⁻¹⁵ to 10⁻¹¹ tesla) without any associated side effects (15). For context, the Earth’s magnetic field ranges from 25 × 10⁻⁶ to 65 × 10⁻⁶ tesla, while cardiac magnetic resonance imaging relies on field strengths of 1.5 to 3.0 tesla (15). Having no side effects, it can be broadly applied including vulnerable populations such as pregnant women (16, 17), patients with renal disease, and children. In contrast to CMR, MCG does not require ECG gating and can yield reliable data even in patients with arrhythmias or elevated heart rates. Operating without physical contact, the system minimizes the risk of cross-contamination and rarely poses a problem for patients with claustrophobia.

Previous clinical research has employed MCG for detecting cardiac arrhythmias in fetuses in utero (17–19) and a substantial body of literature highlights the clinical utility of MCG in the assessment of ischemic cardiomyopathy (20–27). Emerging evidence also suggests that MCG may aid in identifying heart transplant rejection, potentially by detecting myocardial inflammation (28). However, there is only little evidence on its potential to detect non-ischemic cardiomyopathy (15, 29).

In this study, we tested diagnostic accuracy of MCG in detecting non-ischemic cardiomyopathy prospectively, after we established a diagnostic cutoff successfully in a retrospective study (10).

## Methods

We sought to investigate prospectively the diagnostic accuracy of MCG in detecting non-ischemic cardiomyopathy in patients with angina, but non-obstructive coronary artery disease (ANOCA). To rule out obstructive coronary artery disease (CAD) we performed coronary angiography or coronary CT scans. Patients were enrolled after CAD rule-out in the emergency department, in-patient wards and outpatient clinics. Diagnostic accuracy was compared to results obtained by the current gold standards of advanced imaging, i.e. cardiac magnetic resonance (CMR) imaging and/or fludeoxyglucose-18 (FDG-18) positron emission computed tomography (PET-CT) and where available histology and immunohistology from endomyocardial biopsy (EMB)(5, 30). CMR included T1 and T2 maps, additional to late gadolinium enhancement imaging. While CMR served as the first-line advanced imaging modality, PET-CT was performed in patients with recurrent chest pain when prior CMR evaluations were inconclusive (Supplemental Figure 1 and 2). The time window between the MCG measurement and advanced imaging with CMR and/or PET-CT was less than 30 days and there were no changes in therapy between the two measurements. In patients who underwent PET-CT due to inconclusive prior CMRs, the CMR was often performed outside the 30-day time window and thus excluded. For these patients, PET-CT results were used in the study analysis.

Results from EMB took precedence over advanced imaging for the diagnosis. Results from MCG were independently assessed by two trained professionals blinded to the clinical diagnosis, with a third expert to adjudicate in cases of disagreement. Medical data were exported from the electronic medical records at the time of the MCG measurements.

### MCG – Data Acquisition and Measurements

MCG is based on detecting the movement of ions within the myocardium. When an action potential in the heart is generated, it creates voltage changes and consequently an electromagnetic field (10). The system is presented in Figure 1.

**Figure 1.**
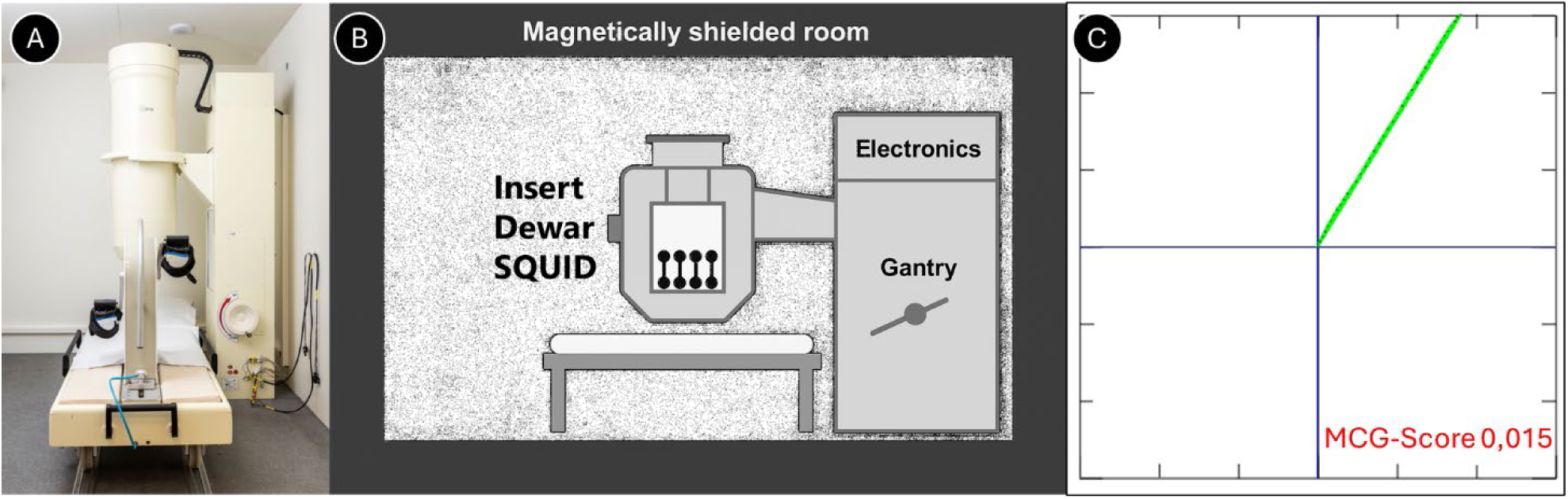
Image of the Magnetocardiography (MCG) System. Panel A shows a photograph of the MCG system used in this study. Panel B provides a schematic illustration of the system layout and setup. Panel C shows an example of a healthy individual with a T-beg–T-max (MCG Score) value of 0.015, which is below the pathological cut-off of >0.051.

The strength and direction of this field are affected by the flow of ions both inside and outside the cells. Typically, the magnetic field produced by the heart ranges from 10^−15^ to 10^−11^ Tesla (10).

The MCG system utilizes an array of 64 highly sensitive magnetic sensors known as superconducting quantum interference devices (SQUIDs, CS-MAG III, Biomagnetik Park GmbH, Hamburg, Germany). These sensors are placed in a shielded environment to reduce interference from external electromagnetic sources. SQUIDs capture variations in the heart’s magnetic field throughout the cardiac cycle and correlate these changes with the QRS complex. To filter out electromagnetic noise, several frequency filters are applied. Measurements provide a three-dimensional view of the magnetic field, which is used to generate a composite vector representing the primary electrical axis of the heart. In assessing non-ischemic cardiomyopathy, the focus is on the vector associated with the T-wave of the action potential, particularly between the beginning of the T-wave on a 12-lead ECG to the maximum of the T-wave (T-beg-Tmax interval). A T-beg-Tmax (MCG Score) value >0.051 has been identified as indicative of pathology, as demonstrated in our previous research (10).

The cohort of the current study consisted of patients who presented to the Department of Cardiology at our university clinic (Charité Universitätsmedizin Berlin) with suspected non-ischemic cardiomyopathy. The inclusion criteria were: Adult patients experiencing symptoms suggestive of non-ischemic cardiomyopathy, including chest pain or pressure, dyspnea, and/or heart racing, after rule out of obstructive coronary artery disease by coronary angiography or coronary CT scans (6). Patients had to be stable enough to undergo advanced imaging and MCG measurements. Patients on immunosuppressive therapy were excluded. Each patient was scanned twice, and the measurement with the better signal-to-noise ratio was used for this study. Signal-to-noise ratio was defined as the number of high-quality QRS complex signals within 60 seconds.

We performed 1:2 propensity score matching based on sex and age decade with healthy controls. Healthy controls were asymptomatic, had no history of cardiac disease, and were unremarkable on physical exam, 12-lead ECG, and echocardiography.

### Power Calculation

A power calculation indicated the need for 110 patients in Cohort 1 with ANOCA, providing an estimated 80% power at a 5% significance level. The prevalence of non-ischemic cardiomyopathy among patients with ANOCA was assumed to be approximately 13%, based on previous findings from our group(31).

### Ethics Committee Approval

This study was reviewed and approved by the ethics committee of DHZC Universitätsmedizin Berlin. All patients provided their written informed consent. The data were analyzed by Phillip Suwalski, and January Weiner. The study was pre-registered on ClinicalTrials.gov ID NCT06689098.

### Statistical analysis

The statistical analyses were performed using RStudio (Posit Software, PBC; Version 2023) and multiple R-packages.

For analysis, age was grouped into decades, with each category representing a 10-year interval (e.g., “10” = ages 10–19, “20” = ages 20–29, etc.). Notably, the decade “10” included only patients aged 18–19 years, as our study only included adults. Appropriate parametric and non-parametric tests including T-tests and the Mann-Whitney U-test were applied as indicated by the data distribution. Results with a p-value < 5% were considered statistically significant.

## Results

### Cohort Description

We enrolled a total of 330 individuals (110 patients and 220 healthy controls) over a two-year period (table 1). As per inclusion criteria, patients presented with symptoms suggestive of non-ischemic cardiomyopathy including chest pain or pressure, dyspnea, and/or palpitation. Of these, 31.8% (n = 35) were women, and the median age of the entire cohort was 43 years (IQR = 24,72, Figure 2). The most common age group among men was 30–39 years and 50-59 years in women.

**Table 1.** illustrates baseline characteristics of the patient and control population. The healthy cohort (Controls) underwent limited clinical evaluation focused on exclusion of comorbidities. Advanced phenotypic characterization was undertaken only in patients with disease. Please see data supplement S1. The abbreviations used are: IQR (interquartile range) and LVEF (left ventricular ejection fraction). * Indicates a significant difference between the groups with a P-value below 0.001.

| Variable |  | Controls | Patients | P-Value |
| --- | --- | --- | --- | --- |
| **Sex** | **Male** | 150 (68.2%) | 75 (68.2%) | 1.000 |
|  | **Female** | 70 (31.8%) | 35 (31.8%) | 1.000 |
| **Age in years (Median, IQR)** |  | 42.500 (24.00) | 43.254 (27.72) | 0.658 |
| **T-beg-Tmax (MCG Score) (Median, IQR)** |  | 0.029 (0.057) | 0.054 (0.052) | < 0.001 * |
| **LVEF (N, % of Group)** | **<30%** | 0 (0.0%) | 15 (13.6%) | < 0.001* |
|  | **>55%** | 220 (100.0%) | 55 (50.0%) | < 0.001* |
|  | **30-40%** | 0 (0.0%) | 10 (9.1%) | < 0.001* |
|  | **40-50%** | 0 (0.0%) | 12 (10.9%) | < 0.001* |
|  | 50-55% | 0 (0.0%) | 18 (16.4%) | < 0.001* |
| Art. Hypertension |  | 21 (9.5%) | 40 (37.0%) | < 0.001* |
| Hypercholesterolemia |  | 1 (0.5%) | 30 (27.8%) | < 0.001* |

**Figure 2.**
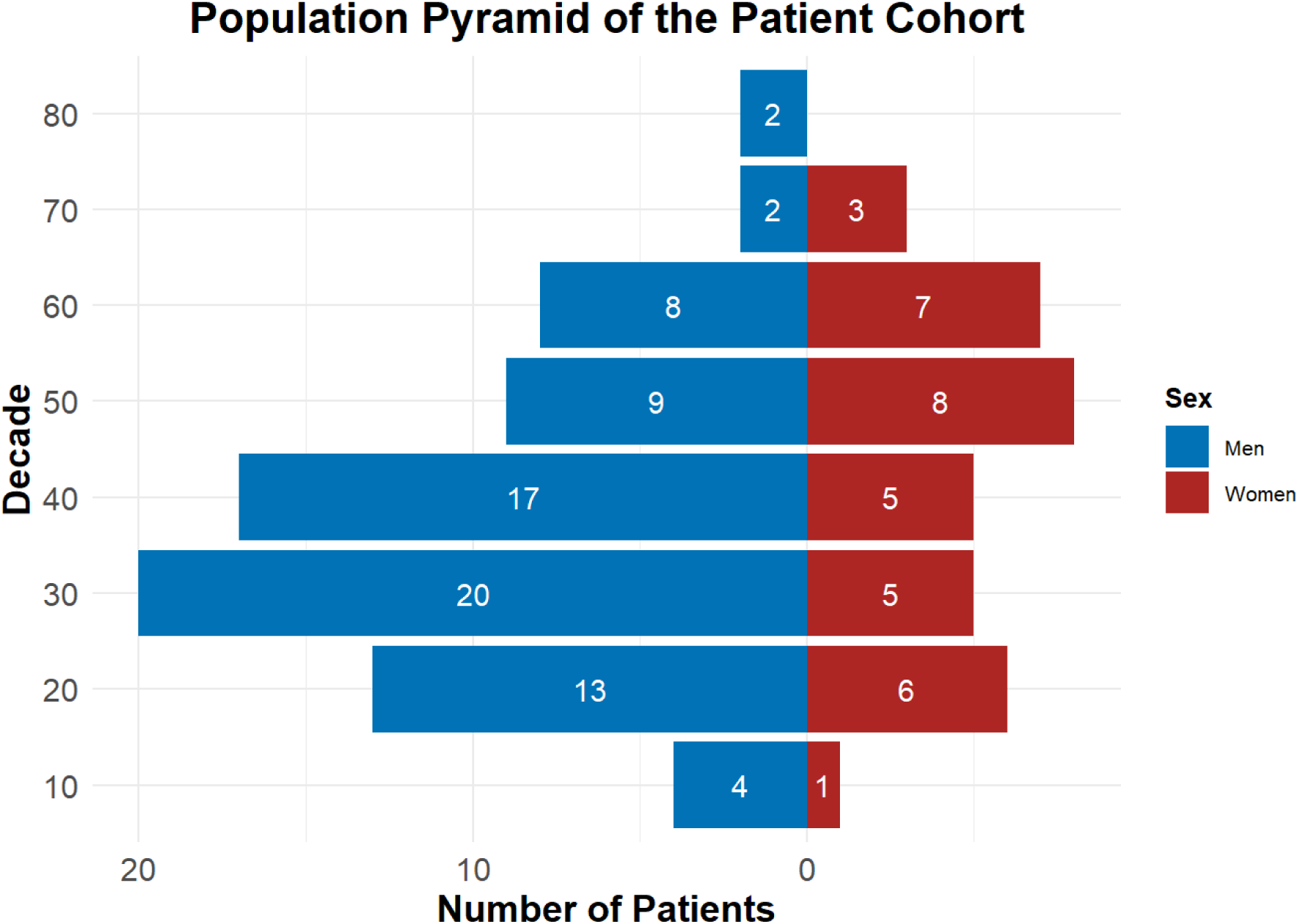
Illustration of the population pyramid of the patient cohort depicting age and sex distribution. Age is illustrated in decades; decade “10” includes only adult patients aged 18–19 years.

The median BMI of the patients was 27,50 kg/m² (IQR = 5,65).

All healthy controls demonstrated an LVEF greater than 56% by design and an interventricular septal diameter (IVSd) of less than 11 mm in women and 13 mm in men.

### Validation of MCG results

All patients (n=110) underwent diagnostic evaluation using at least one of the recommended diagnostic gold-standard methods, i.e. cardiac magnetic resonance (CMR), positron emission tomography-computed tomography (PET-CT), and/or endomyocardial biopsy (EMB). Overall, MCG results were validated by CMR in 59% (n = 65) of patients, by EMB in 21% (n = 23), and by PET-CT in 20% (n = 22, supplemental figure 1). The validation consisted of assessing whether one of the diagnostic methods (CMR, PET-CT, or EMB) confirmed or ruled out the presence of a non-ischemic cardiomyopathy in comparison to the MCG.

A total of 85.5% of patients underwent these diagnostic tests within 14 days of MCG measurement, while 14.5% (n = 16) had their diagnostic tests within 30-day time window.

Three patients were excluded after screening due to unreadable MCG scans. An additional 47 patients were excluded after screening, as they received their validating diagnostic procedure (CMR, PET-CT, or EMB) outside the 30-day cutoff period, which was a pre-registered inclusion criterion.

### Diagnostic Accuracy of Magnetocardiography

The MCG parameter T-beg-Tmax (MCG Score) demonstrated a negative predictive value (NPV) of 93.1% (95% CI: 84-98%) and a positive predictive value (PPV) of 94.34% (95% CI: 84-98%). Sensitivity was 94.74% (95% CI: 85-99%) and specificity 98.54% (95% CI: 96-100%). The presumed disease status based on MCG scans—validated by CMR, PET-CT, and/or EMB—is presented in Figures 3 and 4. The diagnostic classification and its distribution across age and sex are also illustrated in Figure 3 and 4. The AUC was 99,3% (95% CI: 84-98%, supplemental figure 3).

**Figure 3.**
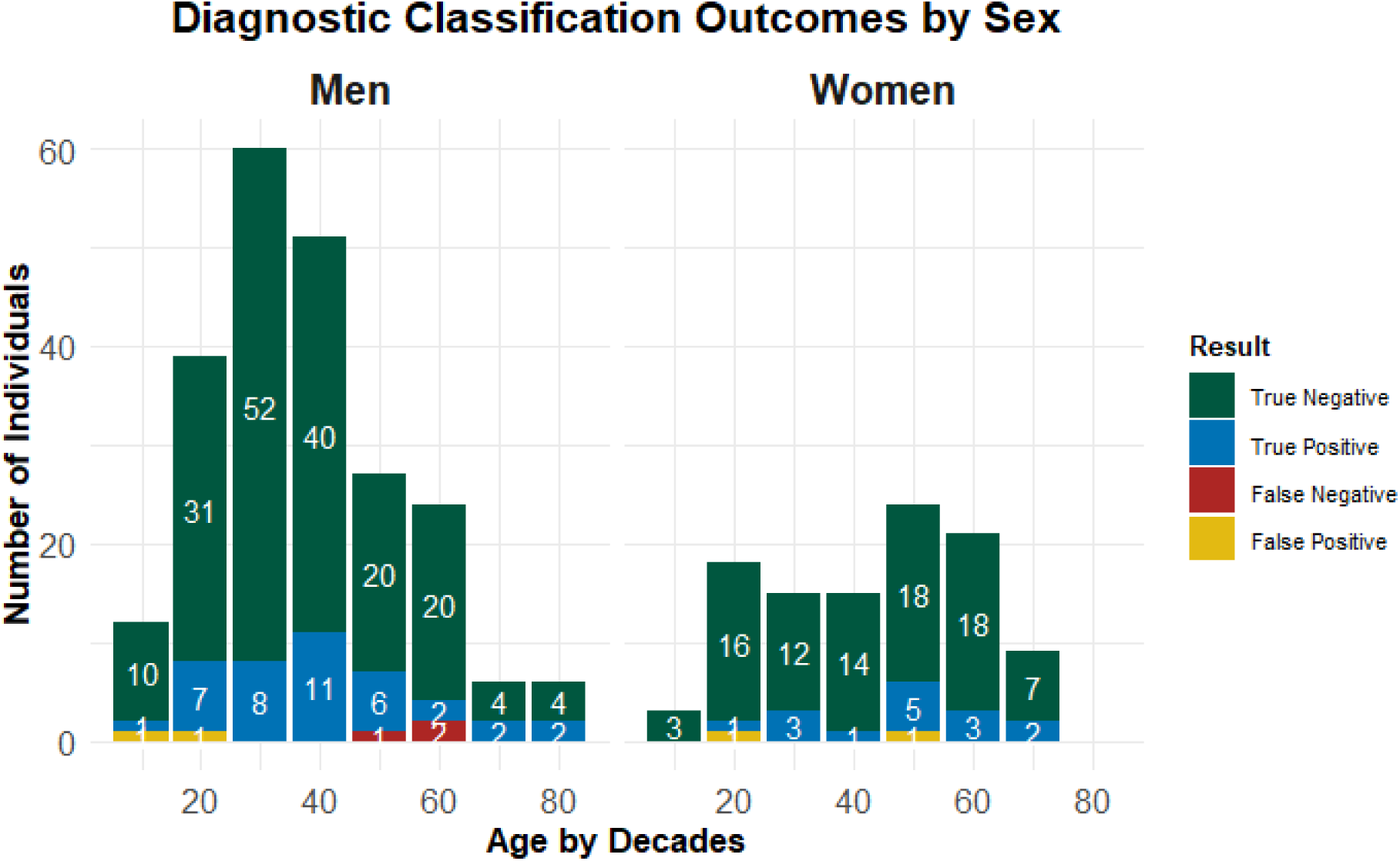
Diagnostic classification outcomes based on sex for 110 patients and 220 healthy controls, matched 2:1 by sex and age decade. The decade “10” in our cohort included only individuals aged 18–19 years, as only adult individuals were enrolled.

**Figure 4.**
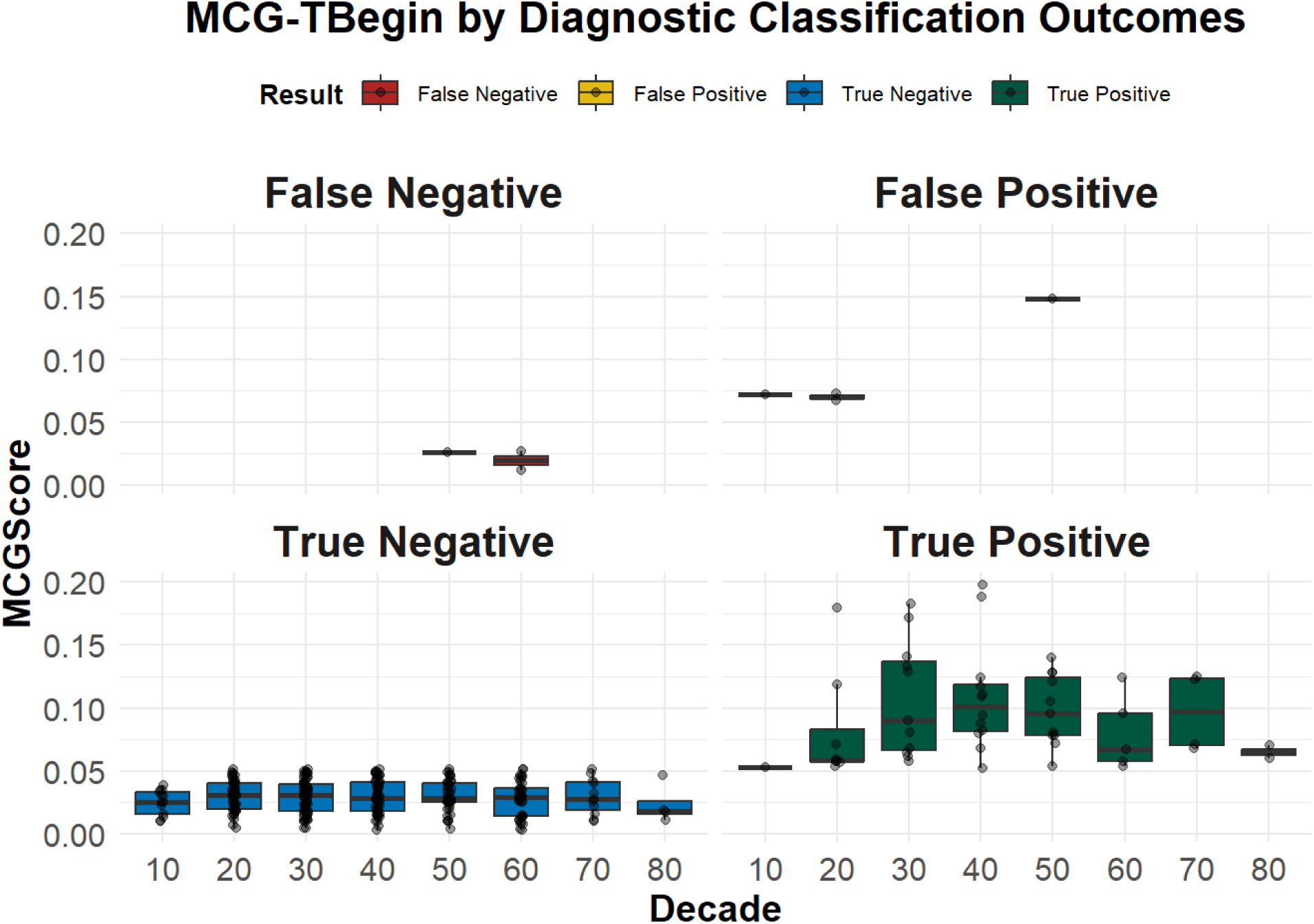
T-beg-Tmax (MCG Score) diagnostic classification based on age decade. The decade “10” in our cohort included only individuals aged 18–19 years, as only adult individuals were enrolled.

The results were highly reproducible (statistical uncertainty: 2.5%, MCG vector +/− 0.004).

The baseline parameters for patients are displayed in supplemental table 1.

### MCG Detection of EMB Proven Inflammatory Cardiomyopathy – Missed by CMR

A total of 30 patients (27%) underwent endomyocardial biopsy in addition to advanced imaging. In three of these patients, CMR failed to detect cardiomyopathy, whereas immunohistology from EMB confirmed inflammatory cardiomyopathy. In all three cases, MCG correctly identified findings consistent with the EMB results (Figure 5). EMBs revealed detection of CD3+ T-cells and CD68+ macrophages indicative of active inflammation. Notably, the patient with the most pronounced inflammatory infiltration on EMB (P3 in Figure 5), characterized by disruption of myocardial architecture by CD3+ T-cells and CD68+ macrophages and fibrosis, also exhibited the most abnormal MCG vector compared to the other two patients (0.128 vs 0.073 and 0.059).

**Figure 5:**
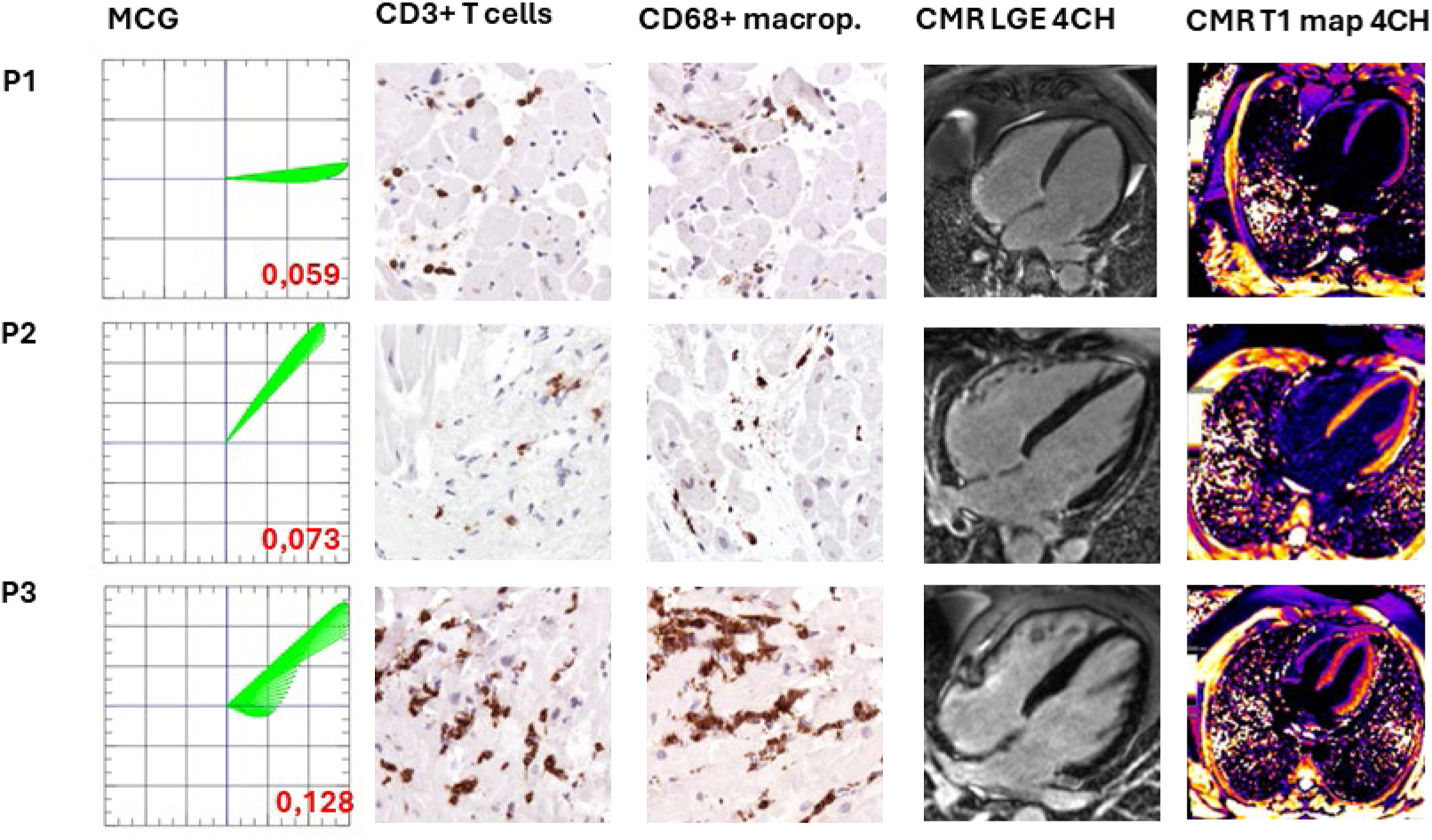
Rows correspond to individual patients —P1: Patient 1, P2: Patient 2, P3: Patient 3. Column 1 displays MCG measurements. Vectors >0.051 are considered pathological; all vectors meet this criterion. Columns 2 and 3 illustrate corresponding endomyocardial biopsy (EMB) samples for P1, P2, and P3. All EMBs are infiltrated by CD3+ T cells and CD68+ macrophages indicative of inflammatory cardiomyopathy. Columns 4 and 5 display corresponding cardiac magnetic resonance imaging (CMR) results for P1, P2, and P3. Utilizing the updated CMR Lake Louise Criteria, no myocardial inflammation was detected.

## Discussion

We demonstrate for the first time in a prospective study that the cardiac magnetic field can be used to detect non-ischemic cardiomyopathies, validating an MCG vector previously derived from retrospective analysis. This MCG vector enables rapid, contactless, and side-effect–free screening that can be deployed broadly, with low threshold to allow early diagnosis and frequent follow-up exams.

To that end, we enrolled 110 patients with symptoms indicative of non-ischemic cardiomyopathy, excluding those with coronary artery disease, and matched them with 220 controls based on sex and age decade. The T-beg-Tmax (MCG Score), validated against current gold-standard diagnostic methods such as CMR, PET-CT, or EMB, demonstrated a remarkable sensitivity of 95% (95% CI: 85-99%), specificity of 99% (95% CI: 96-100%), 93% negative predictive value (NPV, 95% CI: 84-98%) and 94% positive predictive value (PPV, 95% CI: 84-98%).

Given its high diagnostic accuracy, our data suggest that MCG could be a valuable addition to the current diagnostic algorithm for cardiomyopathies, enabling rapid and cost-effective screening that can be broadly implemented in athletes (32), as well as in vulnerable populations such as pregnant women (17, 19), patients with renal impairment, and even in children as a result of the absence of associated adverse effects. Thus, MCG could facilitate the appropriate allocation of advanced imaging to patients who will most likely benefit from such an examination. This is particularly relevant as cardiomyopathies, in particular inflammatory subtypes, remain underdiagnosed, largely due to limited access to advanced imaging or endomyocardial biopsy (2, 8, 33–39).

While the diagnostic sensitivity of CMR in detecting myocarditis varies depending on scar formation and the degree of myocyte necrosis(40, 41), MCG provides a directly measurable parameter, i.e. the T-beg-Tmax (MCG Score), which is unaffected by these potential imaging confounders. By measuring the electromagnetic field of the heart, a result of the flow of ions, MCG is a highly sensitive real time non-invasive diagnostic tool with the ability of detecting even small changes at the molecular level affecting ion flow. This stands in marked contrast to advanced imaging modalities such as CMR, which primarily detect structural features of inflammation. These abnormalities become evident only in the presence of macroscopic tissue involvement and are most reliably visualized several days after the onset of inflammation (42, 43). FDG PET-CT is another valuable method of advanced imaging, which directly reflects changes of metabolism as a result of inflammation, however at a macroscopic level. Moreover, the use of ionizing radiation limits its applicability and raises the threshold for use—particularly in children, pregnant women, and women of childbearing age.

MCG is unlikely to replace established advanced imaging modalities such as CMR or FDG PET–CT, which remain diagnostic gold standards and highly valuable for assessing the extent and pattern of disease involvement (5, 34, 44). Rather, our findings suggest that MCG may serve as a valuable primary screening tool to support clinical decision-making and to guide the judicious use of advanced imaging in patients most likely to benefit.

MCG offers several unique advantages that make it well-suited for broad clinical screening. First, its rapid data acquisition (approximately 1 minute) and fast brief analysis time (approximately 4 minutes) enable high-throughput assessment. Second, the generation of an objective diagnostic score reduces interobserver variability and minimizes training burden required for clinical staff. Third, the absence of adverse effects permits its use across a wide range of patient populations. These features also facilitate serial monitoring, potentially even daily, to guide timely adjustment of treatment algorithms(45). Finally, it provides structured digital output (46), which can be used for artificial intelligence algorithms similar to methods that are currently applied for the ECG (47). In the future, MCG, when combined with structured diagnostic tools such as cardiomyopathy-specific questionnaires (48), could become an important component of routine clinical diagnosis, supporting early detection and targeted evaluation in daily practice (29).

Another potential application of MCG is long-term monitoring of therapeutic response in patients with cardiomyopathies and early detection of recurrence in chronic inflammatory cardiomyopathy (15). This has already been successfully demonstrated in our previous retrospective cohort study (10) and case reports (11, 45, 49). Thus, MCG offers not only a tool for diagnostic assessment but also for treatment monitoring.

### Limitations

A limitation of the current MCG method is the infrastructure required for its operation, including the need for an electromagnetically shielded environment with our current system due to its high sensitivity to magnetic fields in the range of 10⁻¹⁵ Tesla. However, efforts are underway to reduce both complexity and expenses for maintenance, as newer MCG systems are being developed that require weaker or no electromagnetic shielding and no liquid helium (24, 50–52). Whether these systems can reliably detect subtle changes in the electromagnetic field of the heart and therefore be used to identify cardiomyopathy patients and treatment response remains to be determined in future studies. Furthermore, future research should investigate the association between the MCG score and the histological subtypes of nonischemic cardiomyopathies.

A limitation of our present study is that all patients were enrolled in a single center specialized for cardiomyopathies, with a high pre-test probability for the presence of an inflammatory cardiomyopathy in referred patients. We are currently planning a multicenter study to test robustness of the MCG vector further across sites. Patients with severe conditions not stable enough for imaging and MCG measurement could not be examined. Also, patients with intracardiac metallic devices have to be excluded, as such devices are known to interfere with the highly sensitive electromagnetic field measurements of an MCG (10). Currently, no reliable artifact correction is available for patients with pacemakers or implantable cardioverter-defibrillators (ICDs). We are planning to address this limitation in the future through additional research.

## Conclusion

In our study, we enrolled 110 patients with symptoms indicative of non-ischemic cardiomyopathy, excluding those with coronary artery disease, and matched them with 220 controls based on sex and age. The T-beg-Tmax (MCG Score), validated against current gold-standard diagnostic methods such as CMR, PET-CT, and/or endomyocardial biopsy, demonstrated a sensitivity of 94.7% and a specificity of 98.5%.

These findings suggest that MCG could serve as a valuable clinical screening tool for non-ischemic cardiomyopathy in patients with dyspnea, chest pain, or fatigue for eventual referral to advanced imaging.

Implementing MCG into clinical practice may shorten time to diagnosis, improve allocation of personnel and financial resources including the use of advanced imaging and allow for frequent serial assessment and long-term monitoring. Our data indicate that MCG may be an efficient tool for allocation of advanced imaging to those in greatest need.

## Data Availability

An anonymized dataset and code can be made available upon request. Access is only granted to academic institutions and after signing a data sharing agreement.

## Author Contributions

PS, FW, EL, GP, GS and HB enrolled the patients, measured and analyzed the MCG and imaging data. PS provided the extracted EMR data. KK performed the histological analysis of the EMB samples during clinical routine. SK, PS and HB extracted and analyzed the CMR data. PS, JW, DB and HB conducted the statistical analysis. PS, UL, TFL and HB designed the study. HB and PS supervised and were involved in the coordination of all project parts. All authors have written and reviewed the manuscript.

## Acknowledgements

The magnetocardiography device and the service of the machine used in the present study were provided by Biomagnetik Park Holding GmbH. The visual abstract was created in BioRender. Suwalski, P. (2025).

## Supplement

### Figures

**Supplemental Figure 1.**
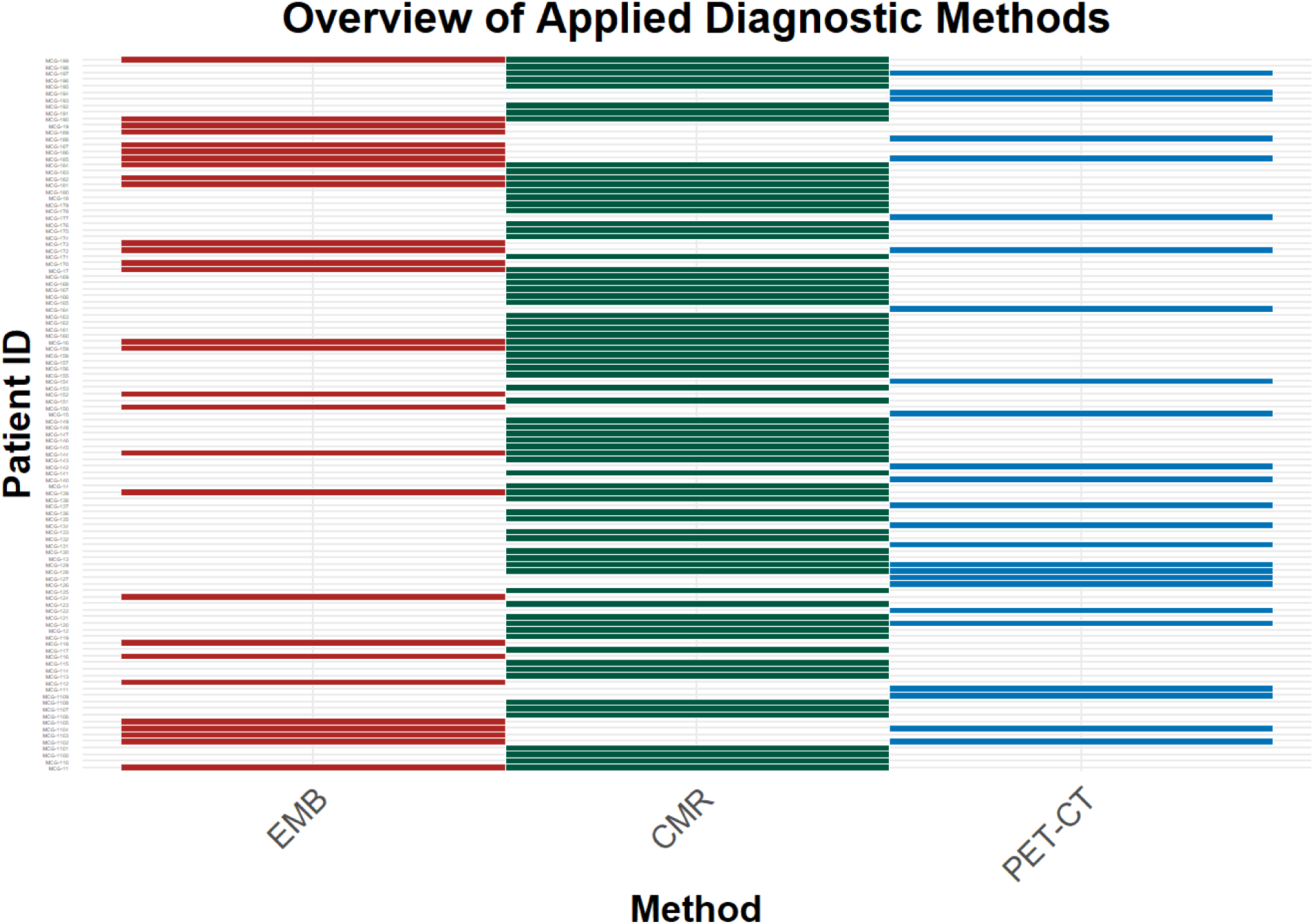
The figure provides an overview of the diagnostic procedures performed in the patient cohort. EMB refers to endomyocardial biopsy, CMR to cardiac magnetic resonance imaging, and PET-CT to positron emission tomography–computed tomography.

Not all diagnostic modalities were available for every patient, as their use was determined by the treating physician based on clinical indication. CMR was the standard imaging modality for the evaluation of cardiomyopathy (5). PET-CT was primarily reserved for the assessment of suspected cardiac sarcoidosis, in particular if CMR was unrevealing (53). EMB was performed in particular in patients with severely reduced left ventricular function (LVEF), in accordance with current expert consensus statements and guidelines (5, 34).

**Supplemental Figure 2.**
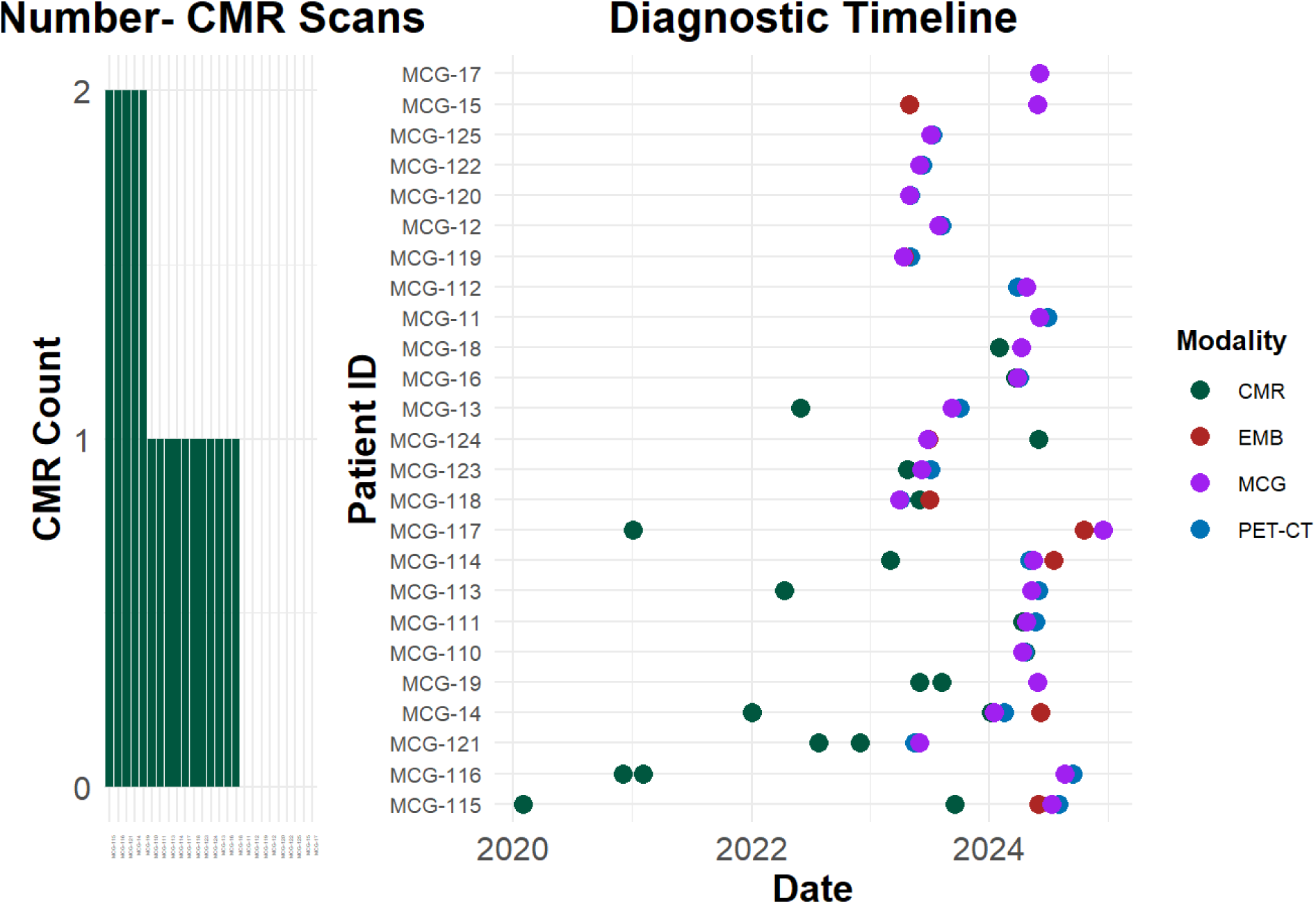
The visualization displays the number of prior cardiac magnetic resonance scans (left panel) and provides an overview of the diagnostic timeline and modalities used to confirm or exclude the diagnosis of cardiomyopathy or cardiac sarcoidosis. EMB refers to endomyocardial biopsy, CMR to cardiac magnetic resonance imaging, MCG to magnetocardiography and PET-CT to positron emission tomography–computed tomography.

Supplemental Figure 2 illustrates, using the subset of patients who underwent PET-CT, the often resource intensive prior diagnostic work-up required to establish a diagnosis. A substantial proportion of these patients had previously been evaluated at external hospitals over the course of several years or months and were referred to our clinic for a second opinion—particularly those with prior CMR studies. At our center, patients underwent CMR, PET-CT, and/or endomyocardial biopsy, as clinically indicated and a magnetocardiography (MCG) study. Given that the MCG was able to detect cardiomyopathy with high accuracy, we suggest that MCG could be a valuable screening tool in the future to allocate advanced diagnostic workup early to those at greatest need.

**Supplemental Figure 3.**
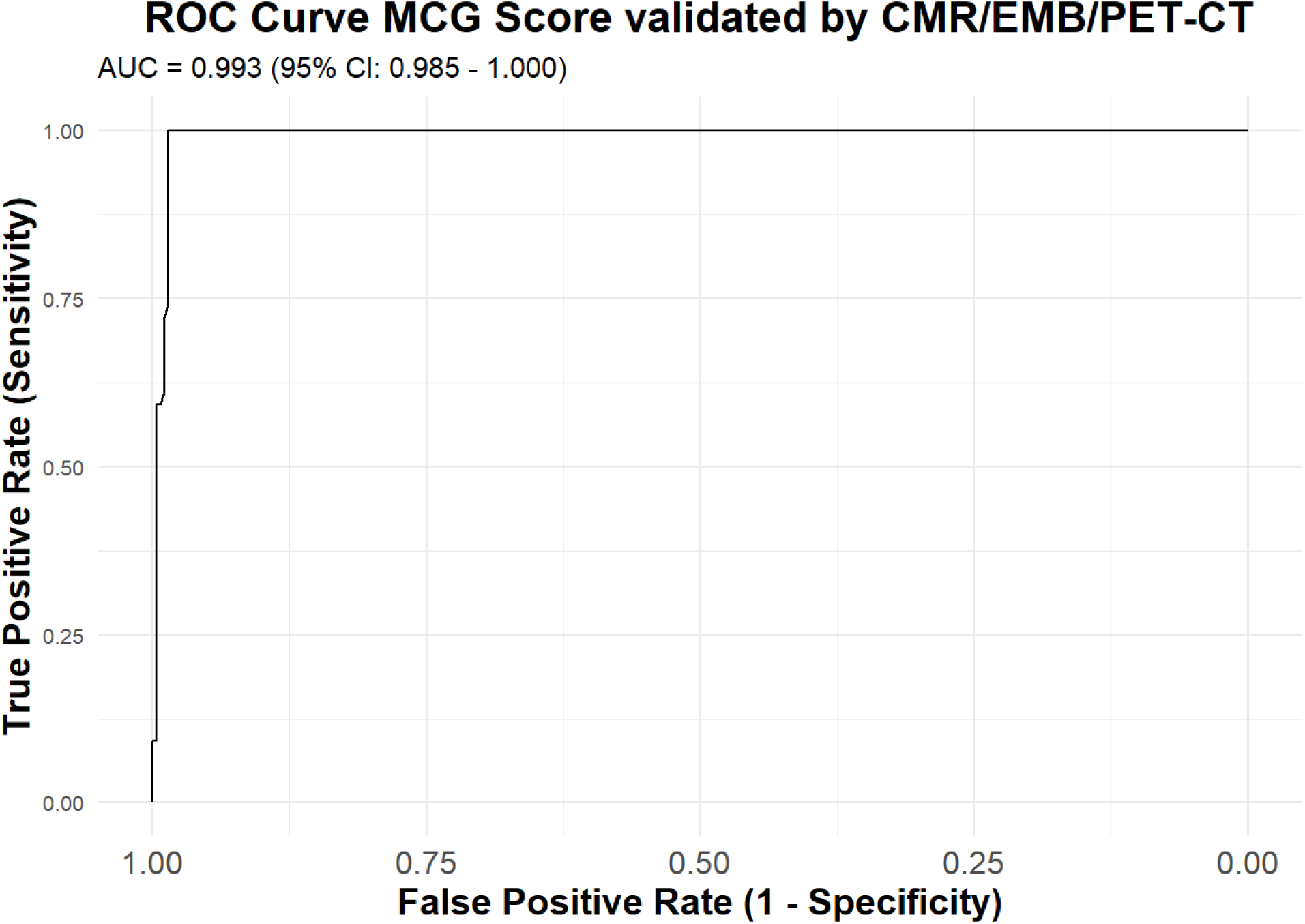
The graph illustrates the area under the curve (AUC) for the magnetocardiography (MCG) score in the present study. Validation of the MCG results took place with cardiac magnetic resonance imaging, positron emission tomography–computed tomography and/or endomyocardial biopsy.

### Tables

**Table S1.** illustrates the baseline characteristics of the patient population divided into categories based on MCG result. Abbreviations: IQR (interquartile range), SGLT2 (sodium-glucose cotransporter-2), MRA (mineralocorticoid receptor antagonist), ACE (angiotensin-converting enzyme) and LVEF (left ventricular ejection fraction). * Indicates a significant difference between the groups with a P-value below 0.05, ** a P-value below 0.01, or *** a P-value below 0.001.

| Variable | True Negative | True Positive | False Negative | False Positive | All | P-Value |
| --- | --- | --- | --- | --- | --- | --- |
| **Sex** |  |  |  |  |  |  |
| **Male** | 31 (63.3%) | 39 (72.2%) | 3 (100.0%) | 2 (50.0%) | 75 (68.2%) | 0.398 |
| **Female** | 18 (36.7%) | 15 (27.8%) | 0 (0.0%) | 2 (50.0%) | 35 (31.8%) |  |
| **Age in years (Median, IQR)** | 38.48 (18.5) | 44.45 (23.38) | 61.45 (1.08) | 25.91 (12) | 43.25 (24.7) | 0.030* |
| **BMI in kg/m<sup>2</sup> (Median, IQR)** | 29.15 (3.3) | 25.10 (5.35) | 29.70 (1) | 28.70 (1) | 27.50 (5.65) |  |
| **T-beg-Tmax (MCG Score) (Median, IQR)** | 0.03 (0.02) | 0.08 (0.06) | 0.02 (0.01) | 0.07 (0.02) | 0.05 (0.052) | <0.001*** |
| **Art. Hypertension** | 17 (34.7%) | 19 (35.2%) | 3 (100.0%) | 1 (25.0%) | 40 (36.4%) | 0.232 |
| **Diabetes Mellitus** | 4 (8.2%) | 12 (22.2%) | 0 (0.0%) | 0 (0.0%) | 16 (14.5%) | 0.268 |
| **Hypercholesterolemia** | 9 (18.4%) | 19 (35.2%) | 1 (33.3%) | 1 (25.0%) | 30 (27.3%) | 0.445 |
| **SGLT2-Inhibitors** | 5 (10.2%) | 20 (37.0%) | 2 (66.7%) | 0 (0.0%) | 27 (24.5%) | 0.009** |
| **Diuretics** | 2 (4.1%) | 19 (35.2%) | 2 (66.7%) | 0 (0.0%) | 23 (20.9%) | <0.001*** |
| **MRA-Antagonists** | 3 (6.1%) | 20 (37.0%) | 2 (66.7%) | 0 (0.0%) | 25 (22.7%) | 0.002 |
| **Sacubitril/Valsartan** | 2 (4.1%) | 19 (35.2%) | 2 (66.7%) | 0 (0.0%) | 23 (20.9%) | <0.001*** |
| **ACE-Inhibitors or Sartans** | 15 (30.6%) | 16 (29.6%) | 1 (33.3%) | 1 (25.0%) | 33 (30.0%) | 0.281 |
| **Betablockers** | 15 (30.6%) | 32 (59.3%) | 3 (100.0%) | 3 (75.0%) | 53 (48.2%) | 0.018* |
| **LVEF (Median, IQR)** | 60.00 (12.75) | 49.00 (30.75) | 38.00 (13.5) | 60.00 (1) | 56.00 (20) | <0.001*** |

